# Glucagon-Like Peptide-1 Receptor Agonist in Large Vessel Occlusion Treated by Reperfusion Therapy: A Phase 2 Randomized Trial

**DOI:** 10.1101/2025.05.04.25326940

**Authors:** Hao Wang, Ho Ko, Thomas Leung, Junzhe Huang, Junjie Sai, Yu Liang, Haipeng Li, Jie Zhang, Qingyang Cao, Wentao Zang, Yinfei Li, Sze Ho Ma, Wai Ting Lui, Joseph Choi, Charlie Chan, Jason Wong, Andrew J. Kwok, Karen Ma, Florence Fan, Anne Chan, Vincent Ip, Howan Leung, Yannie Soo, Ka Tak Wong, Billy Lai, CM Chu, Hanson Leung, Anselm Hui, Tom Cheung, Jill Abrigo, Siu Hung Li, Larry Chan, Jonas Yeung, Sangqi Pan, Terry Yip, LT Lui, Trista Hung, Suk Fung Tsang, Xinyi Leng, Bonnie Lam, Vincent C. T. Mok, Rosa H. M. Chan, Thanh N. Nguyen, Wei Hu, Fengyuan Che, Bonaventure Y. Ip

**Affiliations:** Department of Neurology, Linyi People’s Hospital, Linyi, China; Division of Neurology, Department of Medicine and Therapeutics, Faculty of Medicine, The Chinese University of Hong Kong, HKSAR, China; Li Ka Shing Institute of Health Sciences & Gerald Choa Neuroscience Institute, The Chinese University of Hong Kong, HKSAR, China; Department of Imaging and Interventional Radiology, Faculty of Medicine, The Chinese University of Hong Kong, Hong Kong SAR, China; Department of Medicine and Geriatrics, North District Hospital, HKSAR, China; Department of Medicine and Geriatrics, Alice Ho Miu Ling Nethersole Hospital, HKSAR, China; Department of Electrical Engineering, Faculty of Engineering, City University of Hong Kong, Hong Kong SAR, China; Department of Neurology, Neurosurgery, and Radiology, Boston Medical Center, Boston University Chobanian and Avedisian School of Medicine, USA; Department of Neurology, University of Science and Technology China, Anhui, China; Kwok Tak Seng Centre for Stroke Research and Intervention, The Chinese University of Hong Kong, HKSAR, China

## Abstract

We aimed to determine the effect of semaglutide in patients with acute large vessel occlusion (LVO) receiving endovascular therapy (EVT). In this phase 2, investigator-initiated, multi-center, prospective, randomized, open-label, blinded endpoint trial conducted in China, we recruited patients with disabling LVO undergoing EVT. Patients were randomized (1:1) to semaglutide therapy (0.5 mg subcutaneous semaglutide before and 1 week after EVT) or standard therapy. The primary efficacy outcome was favorable neurological recovery (modified Rankin Scale 0 to 2 at 90 days). The primary safety outcome was a composite of death, malignant brain edema, and intracranial hemorrhage. Between August 2023 and July 2024, 140 patients were randomized to semaglutide (n=69) or standard therapy (n=71). The primary outcome occurred in 39 (56.5%) in the semaglutide group and 39 (54.9%) in the standard therapy group (adjusted RR 1.05 [95%CI: 0.95, 1.15], p=0.37). The primary safety outcome occurred in 16 (23.2%) in the semaglutide group and 17 (23.9%) in the standard therapy group (adjusted RR 0.99 [95%CI: 0.89, 1.11], p=0.89). We observed treatment effect modification by intravenous thrombolysis (IVT) on semaglutide therapy (p_interaction_=0.02); thus we performed the following exploratory analyses: The primary outcome occurred in 22 (64.7%) in the semaglutide group and 15 (44.1%) in the standard therapy group (adjusted RR 1.18 [95%CI: 1.02, 1.36], p=0.02) in the no-IVT stratum (n=68). Outcomes were similar between two groups in the IVT-stratum. This trial suggested semaglutide was safe in patients with LVO and may improve neurological outcome in patients not receiving IVT. These observations should be confirmed in a phase 3 randomized trial (NCT05920889).

## INTRODUCTION

The glucagon-like peptide-1 (GLP-1) receptor is widely expressed in the central nervous system, modulating pathways that regulate a wide range of physiological functions from food intake, glucose homeostasis and metabolism (1, 2), to systemic inflammatory responses (3–5). Neurovascular studies in rodent models demonstrated that GLP-1 receptor agonists (GLP-1RAs) can attenuate neuroinflammatory changes and enhance blood-brain barrier integrity in aging and under ischemic conditions (6–9), as well as counteract reperfusion injury and reduce infarct size following middle cerebral artery (MCA) occlusion (8, 10). These findings suggest that GLP-1RA use during the acute phase may offer neuroprotection, aligning with the Stroke Treatment Academic Industry Roundtable X (STAIR X) recommendation for potential cytoprotective drugs in stroke patients (11).

To date, one phase 2 randomized trial demonstrated that exenatide, a GLP-1RA, did not improve 7-day clinical outcomes in a study population mainly consisting of mild strokes (12). The potential neuroprotective effects of GLP-1RAs remain unexplored in patients with a higher severity of stroke (13). In patients with large vessel occlusion (LVO), successful reperfusion by endovascular therapy (EVT) may enhance the therapeutic potential of GLP-1RAs through two mechanisms: 1) by enabling better drug distribution to vulnerable brain regions with ischemic and reperfusion injury, and 2) by anti-inflammatory effects once blood flow is restored. Therefore, investigating the use of a potent GLP-1RA in the context of LVO strokes reperfused by EVT may uncover the neuroprotective effects of GLP-1RAs in human subjects.

To inform a phase 3 trial, the GALLOP (**G**lucagon-Like Peptide-1 Receptor **A**gonist in **L**arge Vesse**L O**cclusion Stroke Treated by Re**P**erfusion Therapy) randomized trial aimed to determine the safety and signals for efficacy of semaglutide, currently recognized as the most clinically potent GLP-1RA (13), in patients with acute LVO stroke treated by EVT.

## RESULTS

### Trial Population

Between August 11, 2023, and July 25, 2024, 149 patients were screened for study eligibility (**Figure 1**), among which 140 patients were enrolled and randomly assigned to semaglutide (n=69) or standard therapy (n=71). All patients in the semaglutide group received semaglutide injection before puncture, while 3 (4.3%) patients in the semaglutide group did not complete day-7 semaglutide injection due to mortality. The mRS at 90 days was missing for 3 (4.3%) patients in the semaglutide group and 2 (2.8%) patients in the standard therapy group due to loss to follow-up. There was no other missing data. Minor protocol violations occurred in 3 (2.1%) patients (**Table S2**). The trial enrolled to completion. The mean age of patients was 68.2±10.9 years, 94 (67.1%) were male, 46 (32.9%) were female. The median (interquartile range) National Institutes of Health Stroke Scale (NIHSS) was 16 (IQR: 12, 20) and the baseline Alberta Stroke Program Early Computed Tomography Score (ASPECTS) was 8 (IQR: 7, 10). The median time for onset to puncture was 333 (IQR: 194, 440) minutes. Intravenous thrombolysis (IVT), either by alteplase 0.9mg/kg or tenecteplase 0.25mg/kg, was given to 35 (50.7%) in the semaglutide group and 37 (52.1%) in the standard therapy group. No patients received intra-arterial thrombolysis. Baseline characteristics were balanced between the two groups (**Table 1**). All patients were included in the intention-to-treat analyses.

**Figure 1.**
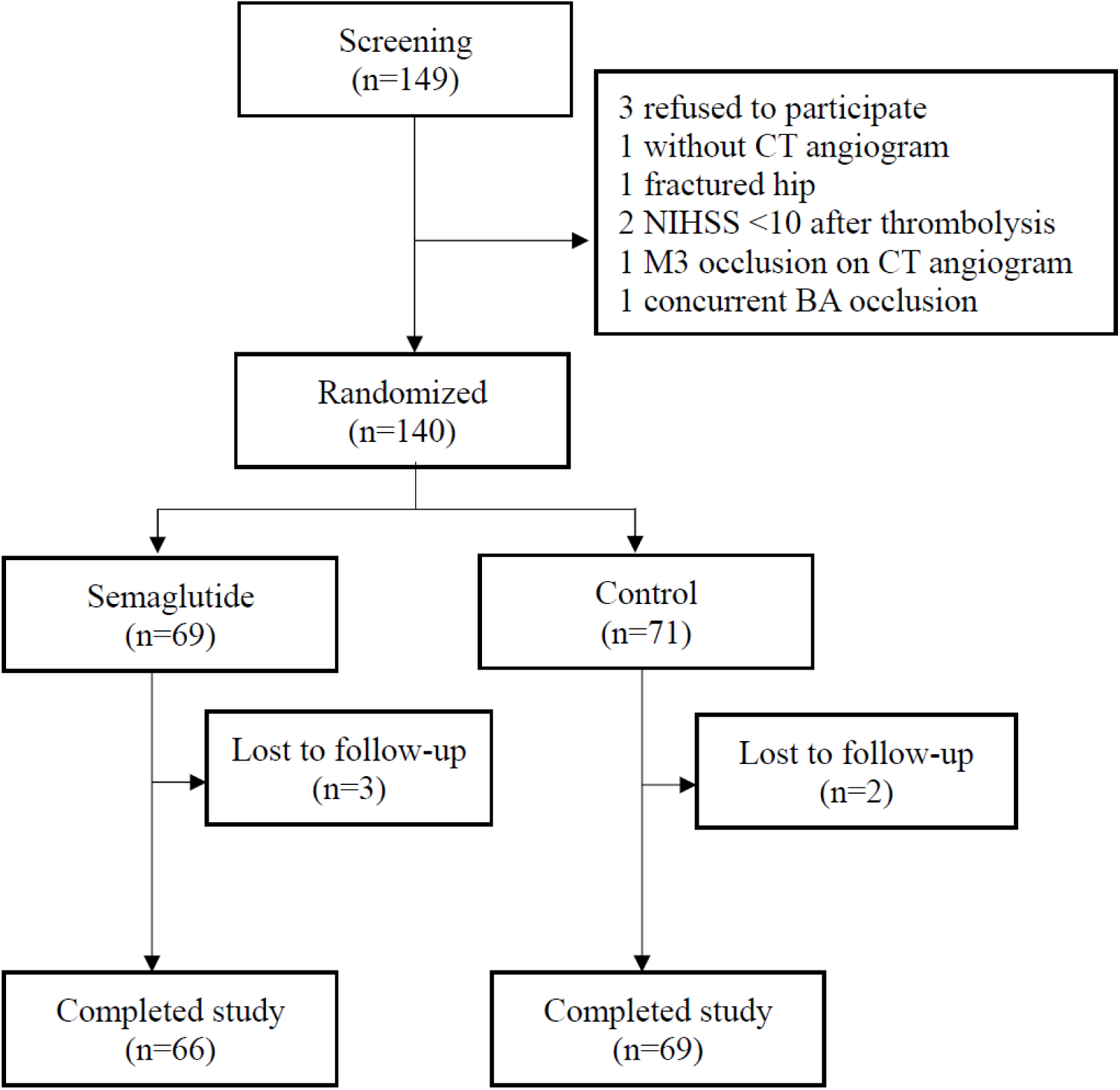
CONSORT flow diagram

**Table 1.** Baseline patient characteristics.

|  | **Semaglutide**<br>**(n=69)** | **Standard**<br>**Therapy (n=71)** | **p-value** |
| --- | --- | --- | --- |
| Age mean(sd) | 69.2 (10.8) | 67.2 (11.1) | 0.26 |
| Male sex n(%) | 44 (63.8) | 50 (70.4) | 0.51 |
| Active smoker n(%) | 17 (24.6) | 16 (22.5) | 0.93 |
| Body weight (kg) mean(sd) | 70.3 (15.1) | 68.2 (12.5) | 0.40 |
| Hypertension n(%) | 42 (60.9) | 48 (67.6) | 0.51 |
| Diabetes mellitus n(%) | 10 (14.5) | 17 (23.9) | 0.23 |
| Atrial fibrillation n(%) | 21 (30.4) | 16 (22.5) | 0.39 |
| Ischemic heart disease n(%) | 11 (15.9) | 5 (7) | 0.17 |
| Congestive heart failure n(%) | 5 (7.2) | 1 (1.4) | 0.11 |
| History of stroke n(%) | 15 (21.7) | 8 (11.3) | 0.15 |
| Peripheral vascular disease n(%) | 1 (1.4) | 1 (1.4) | 1 |
| Chronic kidney disease n(%) | 0 (0) | 2 (2.8) | 0.50 |
| Secondary diversion n(%) | 8 (11.6) | 7 (9.9) | 0.95 |
| Systolic blood pressure mean(sd) | 145.8 (19.6) | 144.3 (23.5) | 0.67 |
| Diastolic blood pressure mean(sd) | 83.5 (11.7) | 85.1 (14.2) | 0.45 |
| Blood glucose on admission (mmol/L) mean(sd) | 7.1 (2.5) | 7.8 (3.2) | 0.15 |
| HbA1c (%) median(IQR) | 5.8 (5.5, 6.1) | 5.9 (5.5, 6.6) | 0.23 |
| ASPECTS median(IQR) | 8.5 (7, 10) | 8 (7, 9) | 0.36 |
| Infarct core (mL) median(IQR) | 9.1 (2.2, 32.3) | 13.2 (3.8, 25.2) | 0.55 |
| Premorbid mRS median(IQR) | 0 (0, 0) | 0 (0, 0) | 0.16 |
| Baseline NIHSS (IQR) | 16 (13, 20) | 16 (12, 20) | 0.39 |
| Intravenous thrombolysis n(%) | 35 (50.7) | 37 (52.1) | 1 |
| Collateral score median(IQR) | 1 (1, 2) | 2 (1, 2) | 0.11 |
| Onset-to-puncture (mins) median(IQR) | 323 (195, 455) | 336 (193, 427) | 0.67 |
| General anesthesia n(%) | 41 (59.4) | 52 (73.2) | 0.12 |
| mTICI 2c or above (%) | 54 (78.3) | 59 (83.1) | 0.61 |
| Aspiration n(%) | 32 (46.4) | 32 (45.1) | 1 |
| Stent retriever n(%) | 8 (11.6) | 6 (8.5) | 0.74 |
| Combined aspiration/stent retriever n(%) | 29 (42) | 33 (46.5) | 0.72 |
| Acute intracranial stenting n(%) | 21 (30.4) | 20 (28.2) | 0.91 |
| **Secondary outcomes** |  |  |  |
| Change between baseline and D3 NIHSS median(IQR) | -8 (-12, -2) | -5 (-9.5, -1.5) | 0.09 |
| Final infarct size (mL) median(IQR) | 13.1 (4.9, 66.4) | 21.8 (10.5, 41.7) | 0.54 |
| Blood glucose on day 3 (mmol/L) mean(sd) | 7 (2.6) | 7.9 (3.1) | 0.09 |
| Change between baseline and D3 glucose (mmol/L) mean(sd) | 0.2 (1.6) | 0.1 (3) | 0.68 |

**Abbreviations:** HbA1c: glycated hemoglobin A1c; ASPECTS: Alberta Stroke Program Early Computed Tomography Score; mTICI: modified Thrombolysis in Cerebral Infarction score; mRS: modified Rankin scale; NIHSS: National Institutes of Health Stroke Scale.

### Primary outcomes

The primary efficacy outcome of modified Rankin scale (mRS) 0–2 at 90 days occurred in 39 (56.5%) in the semaglutide group and 39 (54.9%) in the standard therapy group (adjusted risk ratio [RR] 1.05 [95%CI: 0.95, 1.15], p=0.37, **Figure 2A**). The primary safety outcome, which is a composite of death, malignant edema and intracranial hemorrhage (ICH) occurred in 16 (23.2%) in the semaglutide group and 17 (23.9%) in the standard therapy group (adjusted RR 0.99 [95% CI: 0.89, 1.11], p=0.89).

**Figure 2.**
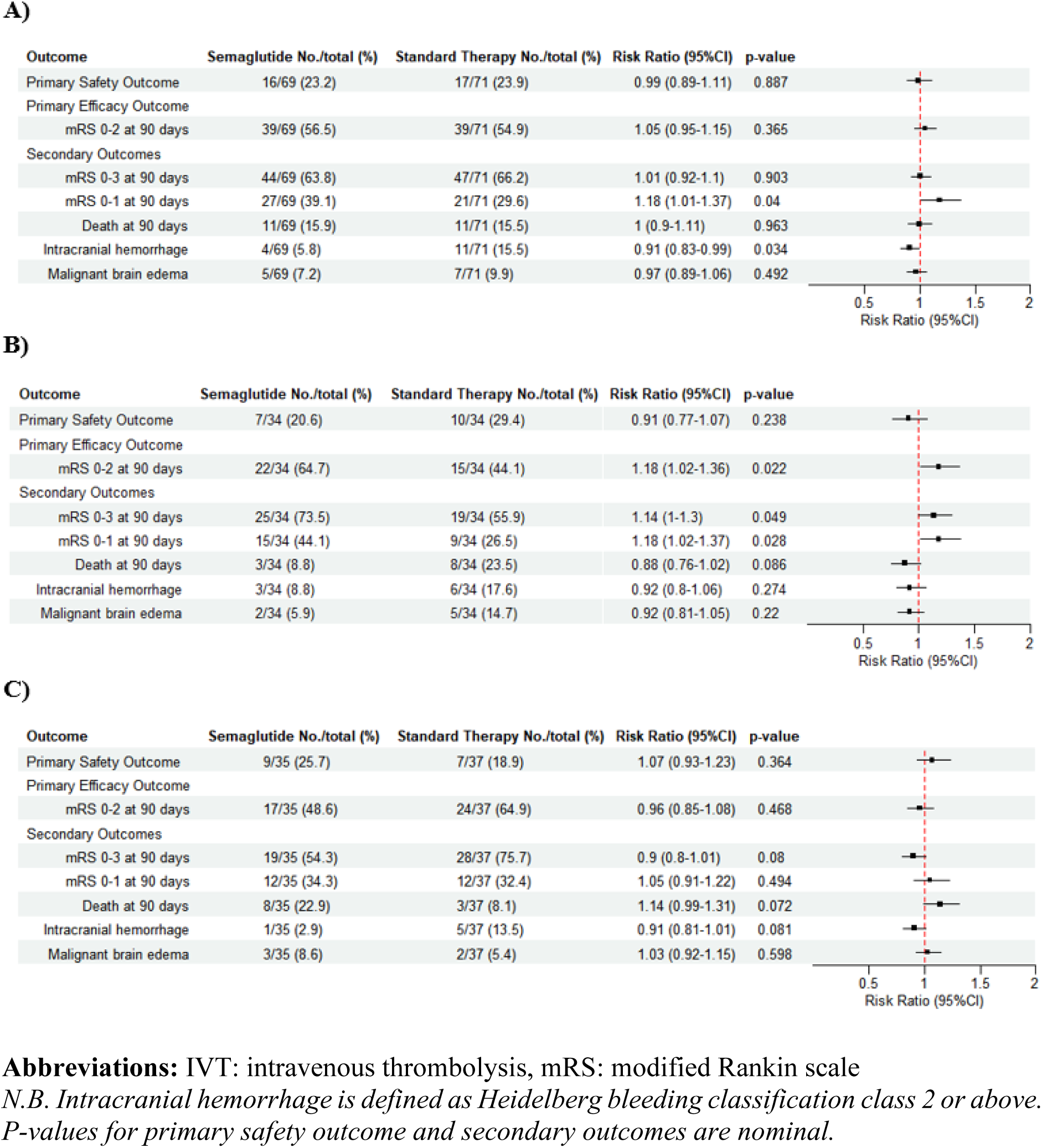
Outcome comparison of A) Overall intention-to-treat sample, B) No-IVT stratum, and C) IVT stratum.

### Secondary outcomes

Secondary outcome comparisons are shown in **Figures 2A-3A**. Semaglutide use was associated with a lower risk of ICH (5.8% vs 15.5%, adjusted RR 0.91 [95%CI: 0.83, 0.99], p=0.03), and a higher likelihood of mRS 0–1 at 90 days (39.1% vs 29.6%, adjusted RR 1.18 [95%CI: 1.01, 1.37], p=0.04). mRS 0–3, death, malignant brain edema and ordinal shift of mRS at 90 days were similar between the two arms. Final infarct sizes were not statistically different between semaglutide and standard therapy (13.1 [IQR: 4.9, 66.4] mL vs 21.8 [IQR: 10.5, 41.7] mL, p=0.58). Unadjusted risk ratios of primary and secondary outcomes are reported in **Table S5**.

**Figure 3.**
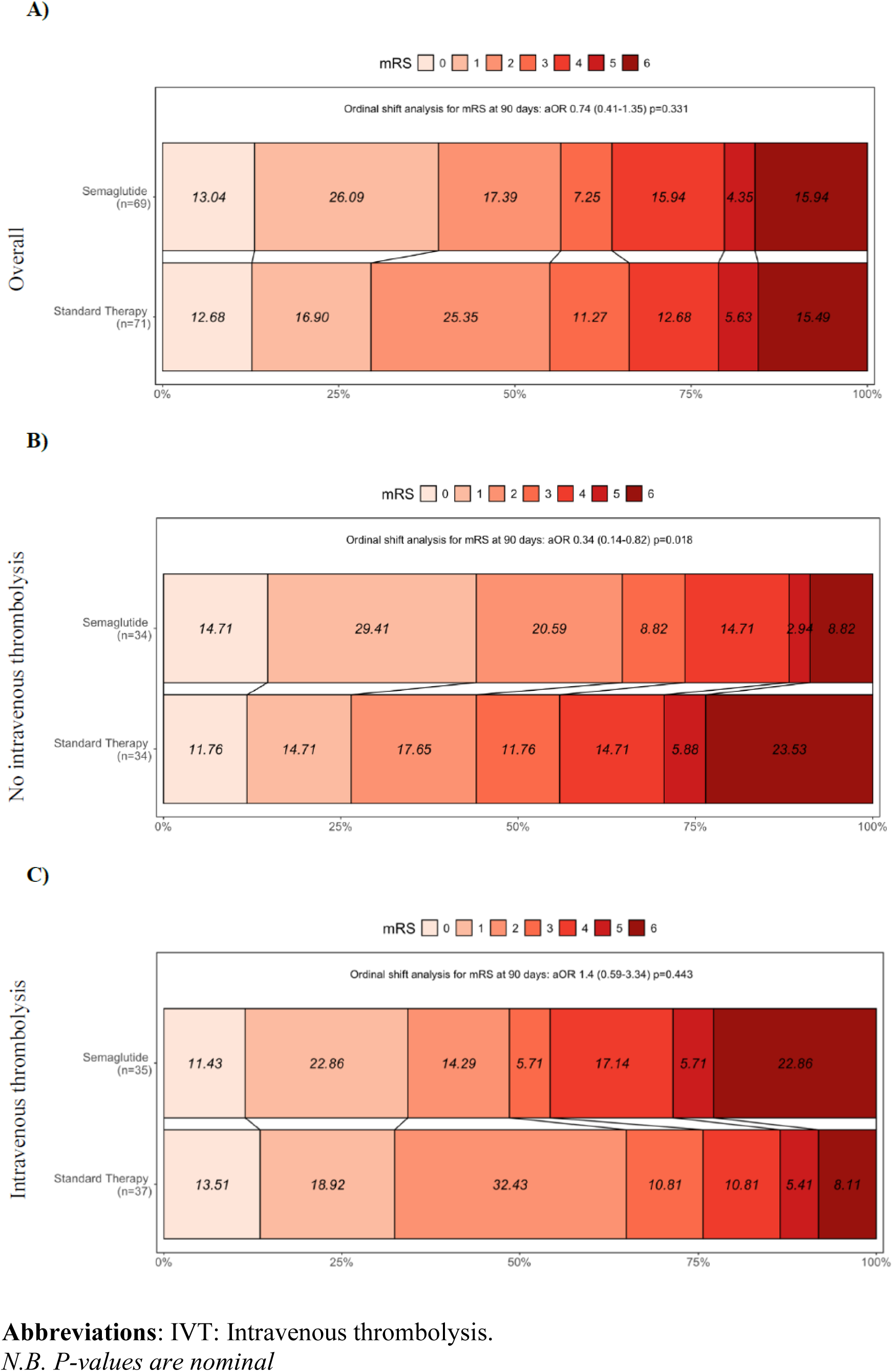
Ordinal shift of modified Rankin scale (mRS) of A) Overall study sample, B) No-IVT stratum, and C) IVT stratum.

### Exploratory analyses

We looked for treatment effect modification by covariates pre-specified in the statistical model. We found that IVT significantly interacted with the treatment effect by semaglutide (p_interaction_=0.02). Other pre-specified covariates in the statistical model, and post-hoc analyses of diabetes mellitus and blood glucose level did not have treatment effect modification with semaglutide.

Exploratory analyses by stratification according to IVT status were subsequently performed. In patients who did not receive IVT, the primary outcome of mRS 0-2 occurred in 22 (64.7%) patients in the semaglutide group (n=34) and 15 (44.1%) patients in the standard therapy group (n=34) (adjusted RR 1.18 [95%CI: 1.02, 1.36], **Figure 2B**). In patients who received IVT, the primary outcome of mRS 0–2 occurred in 17 (48.6%) patients in the semaglutide group and 24 (64.9%) patients in the standard therapy group (adjusted RR 0.96 [95%CI: 0.85, 1.08], **Figure 2C**). Baseline characteristics of both arms in each stratum were similar, except for a higher systolic blood pressure on presentation and lower collateral score among the semaglutide recipients in the IVT stratum (**Tables S3-4**). The onset to puncture time was shorter in the IVT stratum compared to the no-IVT stratum (262 [IQR: 171, 397] minutes vs. 359 [IQR: 248, 521] minutes, p=0.002). The proportional odds ordinal logistic regression suggested treatment effect modification on the ordinal mRS shift by IVT (p_interaction_=0.03, **Figure 3A-C**). There was no violation of the proportional odds assumption. An ordinal shift towards better functional recovery was observed with semaglutide therapy in the no-IVT stratum (adjusted odds ratio 0.34 [95%CI: 0.14, 0.82], **Figure 3B**).

In the no-IVT stratum, we observed a higher proportion of mRS 0–1 at 90 days (44.1% vs 26.5%, adjusted RR 1.18 [95%CI: 1.02, 1.37]), mRS 0–3 at 90 days (73.5% vs 55.9%, adjusted RR 1.14 [95%CI: 1.00, 1.30]), greater reduction in day-3 NIHSS (−8 [IQR: −12, −4] vs −3.5 [IQR: −6, −0.8]), and a lower day-3 blood glucose (6.4±1.4 vs 7.8±3.1) with semaglutide therapy. However, the change between day-3 and baseline blood glucose with semaglutide was modest (−0.4±1.2 mmol/L vs 0.2±2.7 mmol/L). All secondary outcomes were not different between semaglutide and standard therapy in the IVT stratum (**Tables S3-4**).

### Adverse events and sensitivity analysis

The rates of adverse events are listed in **Table 2**. Death occurred in 11 (15.9%) patients in the semaglutide group and 11 (15.4%) patients in the standard therapy group (unadjusted RR 1.00 [95%CI: 0.9, 1.11], p=0.96). Symptomatic ICH, defined as ICH with NIHSS increase of ≥ 4 points, occurred in 1 (1.4%) patient in the semaglutide group and 7 (9.9%) patients in the standard therapy group (unadjusted RR 0.92 [95%CI: 0.85, 0.98], p=0.017). No patients developed hypoglycemia. Complete case analysis showed similar findings (**Tables S6-8**).

**Table 2.** Adverse events.

|  | **Semaglutide (n=69)** | **Standard Therapy (n=71)** |
| --- | --- | --- |
| **Death n(%)** | 11 (15.9) | 11 (15.4) |
| **ICH n(%)** | 4 (5.8) | 11 (15.4) |
| **Symptomatic ICH n(%)** | 1 (1.4) | 7 (9.9) |
| **Malignant brain edema n(%)** | 5 (7.2) | 7 (9.9) |
| **Pneumonia n(%)** | 4 (5.8) | 2 (2.8) |
| **Nausea/Vomiting n(%)** | 3 (4.3) | 1 (1.4) |
| **Hypoglycemia n(%)** | 0 (0) | 0 (0) |
| **Recurrent ischemic stroke n(%)** | 1 (1.4) | 0 (0) |
| **Infection (excluding pneumonia) n(%)** | 3 (4.3) | 1 (1.4) |
| **Hypotension n(%)** | 0 (0) | 1 (1.4) |
| **Euglycemic diabetic ketoacidosis n(%)** | 1 (1.4) | 1 (1.4) |
| **Allergic reactions n(%)** | 0 (0) | 0 (0) |

**Abbreviation**
ICH: intracranial hemorrhage
*N.B. Intracranial hemorrhage (ICH) is defined as Heidelberg bleeding classification class 2 or above. Symptomatic ICH is defined as ICH with increase in National Institute of Health Stroke Scale $\geq 4$ .*

## DISCUSSION

In this phase 2, investigator-initiated, prospective, randomized, open-label, blinded endpoint trial that investigated the use of semaglutide therapy in patients with LVO strokes treated by EVT, semaglutide therapy was well-tolerated without signals of harm. The administration of semaglutide did not delay EVT and was associated with reduced risk of ICH. Moreover, semaglutide showed a trend towards better functional outcome at 90 days, driven by patients who had not received IVT. These findings require confirmation in a larger trial.

Preclinical neuroprotective effects of GLP-1RAs were demonstrated in unilateral transient MCA occlusion animal models, showing reduced infarct volume, oxidative stress, neuroinflammation, neuronal loss and neurological deficit in both diabetic and non-diabetic rodents (14). Notably, the potential neuroprotective effects of GLP-1RA were evident even when drug administration was delayed up to 24 hours following stroke induction (14). Preclinical evidence also suggests that GLP-1RA may enhance blood-brain barrier integrity, which might have reduced ischemic or reperfusion-related brain hemorrhage following EVT as observed in our study (6, 15). Systemically, GLP-1RA may reduce post-stroke stress hyperglycemia, which is a risk factor of blood-brain barrier dysfunction and poor clinical outcomes (16). Clinical evidence exploring the use of GLP-1RA in acute ischemic stroke is scarce. Thus far, only the phase 2 TEXAIS (Treatment with Exenatide in Acute Ischemic Stroke) randomized trial did not demonstrate benefit with exenatide in patients with acute ischemic stroke within 9 hours of onset. The GALLOP trial differs from the TEXAIS in several aspects. First, we included only patients with disabling ischemic stroke. This excluded those with a high likelihood of achieving excellent outcomes, which could have diluted the treatment effects of GLP-1RA due to a floor effect. Second, we focused on patients eligible for EVT. The high rates of excellent angiographic outcomes may have created conditions more similar to those in transient MCA occlusion animal models. Thirdly, we chose semaglutide, which may be more efficacious than exenatide in terms of cardiovascular protection and regulation of systemic stress and inflammatory responses (17, 18), as reflected in post-stroke glucose stabilization.

The treatment effect modification observed with IVT on semaglutide was unexpected. At present, there are no available preclinical or clinical studies that evaluated potential interactions between semaglutide and tenecteplase or alteplase (19). On the one hand, any potential direct interference of GLP-1RA activity by tenecteplase or alteplase, or vice versa, should be ruled out. On the other hand, GLP-1RA may inhibit the expression of plasminogen activator inhibitor-1 (PAI-1) (20, 21), which inhibits tissue plasminogen activators and the conversion of plasminogen to plasmin (22). Downregulation of PAI-1 with co-administration of IVT may potentiate plasmin generation and undesired effects of thrombolytic agents, such as blood-brain barrier disruption (23), brain edema (23), and neurotoxicity (24, 25). The clinical significance of these postulations requires further confirmation. In addition, the interaction between semaglutide and IVT could be confounded by time metrics, as IVT administration is a proxy of early presentation. Since both neuroinflammation and reperfusion injury are time-dependent in ischemic stroke, the therapeutic benefits of GLP-1RAs in modulating or counteracting these processes may only become more apparent beyond the conventional 4.5-hour time window for IVT administration. Further studies are needed to explore whether the potential neuroprotective effect of GLP-1RA is more pronounced in strokes with later presentations.

Our study has several limitations. First, the current phase 2 trial was not powered to draw definite conclusions regarding the efficacy of semaglutide in improving neurological outcomes in patients with LVO, but rather to evaluate the safety and tolerability of semaglutide treatment and estimate any potential treatment effects of semaglutide for a large phase 3 trial. Second, whether similar observations could be reproduced with different dosages of semaglutide require further study. Third, the effect of semaglutide in LVO patients across different ages, LKW-to-presentation time, body weight, ASPECTS, collateral status, blood glucose level on presentation requires further analyses. Last, placebo was not available, but we ensured that all operating neuro-interventionists and raters of functional outcomes were blinded from knowledge of the treatment allocation. All radiological parameters were determined by the central core laboratory with human raters or computer algorithms blinded from treatment allocation and functional outcomes.

In conclusion, 0.5 mg semaglutide administration before and 1 week after EVT in patients with LVO onset within 12 hours was safe and well-tolerated. Semaglutide was associated with lower risk of intracranial hemorrhage and improved neurological outcome in patients who did not receive IVT. These preliminary observations should be confirmed in a large phase 3 trial.

## METHODS

### Study design and participants

The GALLOP trial is a phase 2, investigator-initiated, prospective, randomized, open-label, blinded endpoint (PROBE) trial conducted at four hospitals in China, two of which are thrombectomy centers. We compared the safety and efficacy of semaglutide plus reperfusion therapy, i.e. EVT with or without intravenous thrombolysis (IVT), versus reperfusion therapy alone in patients with disabling LVO. Key inclusion criteria were adults age 18 years or older, LVO at the terminal internal carotid artery (ICA) or M1 segment of the MCA, Alberta Stroke Program Early CT Score (ASPECTS) 6 to 10, National Institutes of Health Stroke Scale (NIHSS) ≥10 at randomization, stroke onset or last-known-well (LKW) ≤12 hours at randomization, and pre-stroke modified Rankin scale (mRS) ≤2. Patients who presented beyond 6 hours from symptom onset or LKW required a CT perfusion scan to assess for clinical-core or perfusion-core mismatch (26, 27). Full inclusion and exclusion criteria are enlisted in **Table S1**. The trial was registered with ClinicalTrials.gov (NCT05920889) and approved by the local institutional review boards (Joint CUHK-NTEC Clinical Research Ethics Committee Reference No.: 2023.026, Science Research Ethics Committee Linyi People’s Hospital Reference No.: YX200651). A detailed study protocol and statistical analysis plan can be found in the **Study Protocol**. The study protocol and statistical analysis plan had been revised once after commencement of study on August 1 2023 (see **Supplementary Note 1**, **Study Endpoints** and **Statistical Analysis**).

### Randomization and masking

All potentially eligible patients underwent computed tomography angiography (CTA) to confirm the LVO. We obtained informed consent from patients or their legal representatives upon confirmation of study eligibility by two investigators. Patients were then randomized in a 1:1 ratio to receive semaglutide plus EVT or EVT alone in the emergency department. Permuted blocked randomization was employed in the study to ensure the balance of subjects throughout the trial setting (28). Randomly generated block sizes of 2, 4, and 6 were adopted to avoid possible mid-block inequality caused by larger blocks. Two blocks with unbalanced treatment distribution were generated at the start and middle of the list. The randomization process was performed using the *blockrand* package (v1.5) in R studio (v4.4.1, R Project for Statistical Computing, RStudio Team 2022). Treatment allocation was concealed until study eligibility was confirmed by two investigators and informed consent had been obtained.

### Procedures

All consecutive patients with LVO who planned to undergo EVT were screened for study eligibility. In addition, study participants underwent screening for intravenous thrombolysis (IVT) by alteplase or tenecteplase before the randomization process. In general, patients with an LKW-to-presentation time of ≤4.5 hours were considered for IVT unless otherwise contraindicated (29). Patients in the semaglutide arm received subcutaneous semaglutide 0.5 mg before arterial puncture and 7 days after EVT. The regimen was chosen based on observational studies that suggested an increase in early blood-brain barrier permeability was associated with worse functional outcomes (30), while a subacute blood-brain barrier permeability increase beyond 10 days after ischemic stroke was associated with good functional outcomes (31). Neurointerventionalists were masked from the treatment allocation. EVT devices were deployed at the discretion of treating interventionists. All study participants were transferred to the stroke unit or neuro-intensive care unit after the procedure, where they received stroke rehabilitation and secondary stroke prevention according to Chinese national guidelines (32).

We prospectively collected patients’ demographic characteristics, medical history, laboratory results and stroke-related parameters including the NIHSS on admission, LKW-to-puncture time and periprocedural details (mode of anesthesia, procedure time, modified Thrombolysis in Cerebral Infarction [mTICI] score). All patients received a follow-up head CT and NIHSS reassessment three days after the procedure or during clinical deterioration to detect any intracranial hemorrhage (ICH) or malignant brain edema (MBE). Brain magnetic resonance imaging (MRI) was obtained 14 to 21 days of randomization for quantification of final infarct size. Clinical follow-up was arranged 90 days after randomization for evaluation of functional recovery by mRS. A telephone or video conferencing follow-up was arranged for patients who were unable to attend an in-person follow-up. All certified assessors for NIHSS and mRS were blinded from the treatment allocation and were not the operating neuro-interventionists of the study subjects.

### Radiological parameters

Images were processed in the central core image processing laboratory. ASPECTS, volumes of infarct core and total ischemic territory on CT perfusion, and final infarct size on MRI brain were determined by in-house automated image-processing algorithms validated with commercially available software. Collateral score (33), mTICI score (34), and hemorrhagic complications were determined by two raters (BI, JA) with more than 10 years of experience (35). Disparities were resolved by a third rater (SM). Raters and computer algorithms were blinded from treatment allocation and clinical outcomes.

### Study Endpoints

The primary efficacy outcome was good neurological recovery, defined as mRS of 0 to 2 at 90 days. The original definition of mRS of 0 to 3 at 90 days was not used after the steering committee discussion on 15 April 2024 as most clinical trials adopted mRS 0 to 2 as the definition for good neurological recovery for anterior circulation LVO (36–38) (**Supplementary Note 1**). The primary safety outcome was a composite of death, MBE (39), and any ICH with Heidelberg Bleeding Classification of 2 or above (35). Secondary outcomes were the ordinal shift of mRS at 90 days, mRS 0 to 3 at 90 days, mRS 0 to 1 at 90 days, death, final infarct size on MRI 14-21 days after randomization, MBE, and ICH. Secondary exploratory outcomes included changes between day-3 and baseline NIHSS and day-3 and baseline blood glucose level. MBE was defined as parenchymal hypodensity of at least 50% of the MCA territory and signs of local brain swelling such as sulcal effacement and compression of the lateral ventricle, and midline shift of ≥5 mm at the septum pellucidum or pineal gland with obliteration of the basal cisterns (39).

### Sample size estimation

No human trials had evaluated the safety and efficacy of GLP-1RA on EVT-eligible patients. We hypothesized that semaglutide had a mild-to-moderate effect size, corresponding to a standardized difference of 0.15-0.25, in reducing poor functional outcome in patients eligible for the study. Considering a rate of 2.5% suboptimal scan qualities, 5% of loss-to-follow-up and 10% of suboptimal recanalization according to the track records of participating centers, 140 participants were required for 90% power and significance level of 0.05 for the main trial (40, 41). Details of the sample size estimation are described in the

### Study Protocol

#### Statistical Analyses

We expressed normally distributed continuous variables as mean ± standard deviation and non-normally distributed continuous variables as median (interquartile range [IQR]). Categorical variables were expressed as number (percentage). We compared baseline continuous variables of semaglutide versus standard therapy by independent sample t-test or Wilcoxon rank-sum test, and categorical variables by Chi-squared or Fisher’s exact test as appropriate.

We used the modified Poisson regression models to evaluate the risk ratios (RR) of the primary and binary secondary outcomes of the two treatment (42). The original plan of multivariable logistic regression was not used as modified Poisson regression could provide unbiased estimation of risk ratios in the setting of common event occurrence compared to logistic regression (**Supplementary Note 1**) (43). Proportional odds ordinal logistic regression was used to compare the ordinal shift of mRS between the two groups. The proportional odds assumption was tested with Brant test. Cox regression was used to compare mortality at 90 days. Analysis of covariance (ANCOVA) was used to compare continuous outcomes. Comparisons of primary outcomes, ordinal shift of mRS, mRS 0-3 and mRS 0-1 at 90 days were adjusted for a pre-specified set of covariates, including age, premorbid mRS, IVT status, NIHSS on presentation, LKW-to-puncture time, mTICI, and baseline ASPECTS. Comparisons of other secondary outcomes were adjusted for NIHSS on presentation. All analyses were performed for the intention-to-treat population, defined as patients who underwent randomization, regardless of treatment received. Deceased patients were considered having an NIHSS of 42. Missing primary outcomes were imputed as the worst possible score for an outcome measure. Complete case analyses for the outcomes were performed in patients without missing data. We performed post-hoc analyses by stratifying IVT status due to the evidence of treatment effect modification by IVT treatment on semaglutide therapy. All secondary outcomes and post-hoc analyses should be considered exploratory.

## Data Availability

Anonymized data will be made available by request to the corresponding authors from any qualified investigator.

## Supporting information

Study protocol

## Data Availability

All data produced in the present study are available upon reasonable request to the corresponding author.

## Non-author contributions to Data collection, Analysis, or Writing/editing assistance

The authors thank the contributions from Dr. Robert Dan (MBBS, The Chinese University of Hong Kong), Mrs. Sophia Dan (BBS, MH, JP, The Chinese University of Hong Kong) and Ms. Anki Miu (MSc, The Chinese University of Hong Kong). Dr. Robert Dan provided general advice for the study. Mrs Sophia Dan provided general advice for the study. Ms Anki Miu provided clerical assistance for the study.

## Access to data and data analysis

Bonaventure Y. Ip had full access to all the data in the study and takes responsibility for the integrity of the data and the accuracy of the data analysis.

## AUTHOR CONTRIBUTIONS

H. W. conducted the study, collected the data, and critically revised the manuscript for intellectual content. H. K. created the study concept and design and supervised the study. T. L. conducted the study, collected the data, and critically revised the manuscript for intellectual content. J. H. conducted the study and collected the data. J. S. conducted the study. Y. Liang conducted the study and collected the data. H. Li verified the data. J. Z. conducted the study and collected the data. Q. C. conducted the study and collected the data. W. Z. conducted the study and collected the data. Y. Li conducted the study and collected the data. S. H. M. conducted the study and collected the data. W. T. L. conducted the study and collected the data. J. C. conducted the study and collected the data. C. Chan conducted the study and collected the data. Jason Wong conducted the study and collected the data. A. J. K. conducted the study and collected the data. K. M. conducted the study and collected the data. Florence Fan conducted the study and collected the data. A. C. conducted the study and collected the data. Vincent Ip conducted the study and collected the data. H. Leung conducted the study and collected the data. K. T. W. conducted the study and collected the data. B. Lai conducted the study and collected the data. C. Chu conducted the study and collected the data. H. Leung conducted the study and collected the data. A. H. conducted the study and collected the data. T. C. conducted the study and collected the data. J. A. conducted the study and collected the data. S. H. L. conducted the study and collected the data. L. C. conducted the study and collected the data. Jonas Yeung conducted the study and collected the data. S. P. did the statistical analysis. T. Y. did the statistical analysis. L. L. critically revised the manuscript for intellectual content. T. H. conducted the study and collected the data. S. F. T. conducted the study and collected the data. X. L. critically revised the manuscript for intellectual content. B. Lam critically revised the manuscript for intellectual content. V.C.T. M. critically revised the manuscript for intellectual content. R. H. M. C. critically revised the manuscript for intellectual content. T. N. N. critically revised the manuscript for intellectual content. W. H. critically revised the manuscript for intellectual content. F. C. critically revised the manuscript for intellectual content. B. Y. I. created the study concept and design, supervised the study, critically revised the manuscript for intellectual content, and verified the data.

