## Supplementary material for "Glucagon-Like Peptide-1 Receptor Agonist in Large Vessel Occlusion Treated by Reperfusion Therapy: A Phase 2 Randomized Trial"

**Title:**

**Table S1. Inclusion and exclusion criteria**

| |
|---|
| **Inclusion criteria** |
| LVO stroke at terminal ICA or proximal M1 eligible for emergency endovascular treatment as per current treatment guideline |
| LKW-to-puncture time ≤ 12 hours |
| Age 18 years or greater |
| National Institute of Health Stroke Scale ≥ 10 |
| LVO stroke due to thromboembolism or intracranial stenosis (acute or acute on chronic occlusion) |
| Patients who received computer tomographic angiography and perfusion |
| Pre-stroke (24 hours prior to stroke onset) independent functional status with modified Rankin Scale ≤ 2 |
| Consent process completed as per national laws and regulation and the applicable ethics committee requirements |
| **Exclusion criteria** |
| ASPECT score ≤ 5 |
| Intracranial hemorrhage on pre-EVT imaging |
| EVT completed before randomization |
| LVO etiologies other than thromboembolism or intracranial stenosis (acute or acute on chronic total occlusion), e.g. arterial dissection, infective endocarditis on initial diagnostic imaging. |
| Estimated or known body mass index < 18 $kg/m^2$ |
| Pregnancy/Lactation; female, with positive urine or serum beta human chorionic gonadotropin (β-hCG) test, or breastfeeding. |
| Creatinine clearance < 30mL/min |
| Severe or fatal comorbid illness, e.g. terminal malignancy |
| Participation in another clinical trial investigating a drug, medical device, or a medical procedure in the 30 days preceding trial inclusion. |
| History of allergy to GLP-1RA |
| Family or personal history of multiple endocrine neoplasia, medullary thyroid carcinoma, pancreatic carcinoma, known proliferative diabetic retinopathy |
| Active sepsis on randomization |
| Patients with hypoglycaemia on presentation. Defined as capillary or serum glucose level of <4mmol/L. |
| Patient already on GLP-1RA prior to screening. |
| Contraindications to iodine-based CT contrast. |

**Abbreviation:** LVO: large vessel occlusion; ICA: internal carotid artery; LKW: last-known-well; ASPECT: Alberta Stroke Program Early Computer Tomography Score; GLP-1RA: glucagon-like peptide-1 receptor agonist; CT: computer tomography.

**Table S2. Details of Protocol Deviation**

| Subject no. | Randomization arm | Details of protocol deviation |
|---|---|---|
| 004 | Standard therapy | NIHSS 9 on presentation |
| 007 | Semaglutide | Onset-to-puncture time 18 hours |
| 013 | Semaglutide | Premorbid mRS 3 |

**Abbreviation:** NIHSS: National Institute of Health Stroke Scale, mRS: modified Rankin scale

**Table S3. Baseline characteristics of the no-IVT stratum**

| Parameters | Semaglutide (n=34) | Standard Therapy (n=34) | p-value |
|---|---|---|---|
| Age mean(sd) | 66.6 (11.8) | 65 (10.5) | 0.56 |
| Male sex n(%) | 22 (64.7) | 22 (64.7) | 1 |
| Active smoker n(%) | 5 (14.7) | 6 (17.6) | 1 |
| Body weight (kg) mean(sd) | 66.6 (13.6) | 69 (12.5) | 0.48 |
| Hypertension n(%) | 21 (61.8) | 23 (67.6) | 0.80 |
| Diabetes mellitus n(%) | 5 (14.7) | 6 (17.6) | 1 |
| Atrial fibrillation n(%) | 14 (41.2) | 5 (14.7) | 0.03 |
| Ischemic heart disease n(%) | 5 (14.7) | 3 (8.8) | 0.71 |
| Congestive heart failure n(%) | 3 (8.8) | 0 (0) | 0.24 |
| History of stroke n(%) | 9 (26.5) | 6 (17.6) | 0.56 |
| Peripheral vascular disease n(%) | 1 (2.9) | 0 (0) | NA |
| Chronic kidney disease n(%) | 0 (0) | 0 (0) | NA |
| Secondary diversion n(%) | 0 (0) | 0 (0) | NA |
| Systolic blood pressure mean(sd) | 140.8 (15.9) | 148.8 (21.3) | 0.08 |
| Diastolic blood pressure mean(sd) | 80.4 (12.3) | 85.8 (13.9) | 0.10 |
| Blood glucose on admission (mmol/L) mean(sd) | 6.7 (1.6) | 7.6 (3.1) | 0.17 |
| HbA1c (%) median(IQR) | 5.7 (5.4, 6) | 5.8 (5.5-6.1) | 0.33 |
| ASPECTS median(IQR) | 8 (7-10) | 8 (6.5, 9) | 0.19 |
| Infarct core (mL) median(IQR) | 8 (1.5, 20.9) | 14.7 (0.8, 23.2) | 0.17 |
| Premorbid mRS median(IQR) | 0 (0, 0) | 0 (0, 0) | 0.57 |
| Baseline NIHSS (IQR) | 16 (11, 20.5) | 16 (10.2, 18.8) | 0.50 |
| Collateral score median(IQR) | 1 (1, 2) | 2 (1, 2) | 0.07 |
| Onset, to, puncture (mins) median(IQR) | 367.2 (256.8, 519) | 354 (242.5, 501) | 0.87 |
| General anesthesia | 18 (52.9) | 26 (76.5) | 0.08 |
| mTICI 2c or above (%) | 29 (85.3) | 29 (85.3) | 1 |
| Aspiration n(%) | 17 (50) | 12 (35.3) | 0.33 |
| Stent retriever n(%) | 2 (5.9) | 3 (8.8) | 1 |
| Combined aspiration/stent retriever n(%) | 15 (44.1) | 19 (55.9) | 0.47 |
| Acute intracranial stenting n(%) | 8 (23.5) | 10 (29.4) | 0.78 |
| **Secondary outcomes** | | | |
| Change between baseline and D3 NIHSS median(IQR) | -8 (-12, -4) | -3.5 (-6, 0.8) | <0.001 |
| Final infarct size (mL) median(IQR) | 11.1 (5.1, 36) | 17.1 (9.8, 25.9) | 0.37 |
| Blood glucose on day 3 (mmol/L) mean(sd) | 6.4 (1.4) | 7.8 (3.1) | 0.02 |
| Change between baseline and D3 glucose (mmol/L) mean(sd) | -0.4 (1.2) | 0.2 (2.7) | 0.06 |

**Abbreviation:** HbA1c: glycated hemoglobin A1c; ASPECTS: Alberta Stroke Program Early Computer Tomography Score; mTICI: modified Thrombolysis in Cerebral Infarction score; mRS: modified Rankin scale; NIHSS: National Institute of Health Stroke Scale.

**Table S4. Baseline characteristics of the IVT stratum**

| Parameters | Semaglutide (n=35) | Standard Therapy( n=37) | p-value |
|---|---|---|---|
| Age mean(sd) | 71.9 (9) | 69.2 (11.4) | 0.27 |
| Male sex n(%) | 22 (62.9) | 28 (75.7) | 0.36 |
| Active smoker n(%) | 12 (34.3) | 10 (27) | 0.68 |
| Body weight (kg) mean(sd) | 73.5 (15.7) | 67.6 (12.6) | 0.09 |
| Hypertension n(%) | 21 (60) | 25 (67.6) | 0.67 |
| Diabetes mellitus n(%) | 5 (14.3) | 11 (29.7) | 0.20 |
| Atrial fibrillation n(%) | 7 (20) | 11 (29.7) | 0.50 |
| Ischemic heart disease n(%) | 6 (17.1) | 2 (5.4) | 0.15 |
| Congestive heart failure n(%) | 2 (5.7) | 1 (2.7) | 0.61 |
| History of stroke n(%) | 6 (17.1) | 2 (5.4) | 0.15 |
| Peripheral vascular disease n(%) | 0 (0) | 1 (2.7) | NA |
| Chronic kidney disease n(%) | 0 (0) | 2 (5.4) | NA |
| Secondary diversion n(%) | 8 (22.9) | 7 (18.9) | 0.90 |
| Systolic blood pressure mean(sd) | 152.6 (19.1) | 140.1 (25) | 0.01 |
| Diastolic blood pressure mean(sd) | 86.4 (10.4) | 84.5 (14.6) | 0.52 |
| Blood glucose on admission (mmol/L) mean(sd) | 7.4 (3.1) | 7.9 (3.3) | 0.49 |
| HbA1c (%) median(IQR) | 6 (5.6, 6.4) | 6.1 (5.7, 6.6) | 0.36 |
| ASPECTS median(IQR) | 8.5 (7, 9.2) | 9 (7, 9.5) | 0.62 |
| Infarct core (mL) median(IQR) | 13.9 (4.4, 44.9) | 12 (4.5, 26.2) | 0.37 |
| Premorbid mRS median(IQR) | 0 (0, 0) | 0 (0, 0) | 0.15 |
| Baseline NIHSS (IQR) | 16 (14, 19.5) | 15 (13, 21) | 0.56 |
| Collateral score median(IQR) | 1 (1, 2) | 2 (1, 2) | 0.03 |
| Onset-to-puncture (mins) median(IQR) | 263 (178, 419.5) | 262 (155, 384) | 0.61 |
| General anethesia n(%) | 23 (65.7) | 26 (70.3) | 0.87 |
| mTICI 2c or above (%) | 25 (71.4) | 30 (81.1) | 0.49 |
| Aspiration n(%) | 15 (42.9) | 20 (54.1) | 0.48 |
| Stent retriever n(%) | 6 (17.1) | 3 (8.1) | 0.30 |
| Combined aspiration/stent retriever n(%) | 14 (40) | 14 (37.8) | 1 |
| Acute intracranial stenting n(%) | 13 (37.1) | 10 (27) | 0.51 |
| **Secondary outcomes** | | | |
| Change between baseline and D3 NIHSS median(IQR) | -8 (-12, 0) | -6 (-11, -3) | 0.75 |
| Final infarct size (mL) median(IQR) | 25.9 (3.3, 122.5) | 28.6 (11.3, 57.2) | 0.28 |
| Blood glucose on day 3 (mmol/L) mean(sd) | 7.7 (3.2) | 8 (3.2) | 0.69 |
| Change between baseline and D3 glucose (mmol/L) mean(sd) | 0.7 (1.8) | -0.1 (3.3) | 0.25 |

**Abbreviations:** HbA1c: glycated hemoglobin A1c; ASPECTS: Alberta Stroke Program Early Computer Tomography Score; mTICI: modified Thrombolysis in Cerebral Infarction score; mRS: modified Rankin scale; NIHSS: National Institute of Health Stroke Scale.

**Table S5. Unadjusted Risk Ratios**

| | **Unadjusted RR (95% CI)** | **p-value** |
|---|---|---|
| Primary safety outcome | 0.99 (0.89, 1.11) | 0.89 |
| mRS 0-2 at 90 days | 1.02 (0.92, 1.13) | 0.74 |
| mRS 0-3 at 90 days | 0.99 (0.90, 1.09) | 0.89 |
| mRS 0-1 at 90 days | 1.09 (0.97, 1.22) | 0.13 |
| Death | 1 (0.9, 1.11) | 0.94 |
| Intracranial hemorrhage | 0.92 (0.83, 1.00) | 0.05 |
| Malignant brain edema | 0.98 (0.90, 1.06) | 0.58 |

**Abbreviation:** mRS: modified Rankin scale, RR: risk ratio

*N.B. Intracranial hemorrhage is defined as Heidelberg bleeding classification class 2 or above.*

**Table S6. Complete case analysis (overall study sample)**

| Parameters | Semaglutide (n=66) | Standard Therapy (n=69) | RR (95% CI) | p-value |
|---|---|---|---|---|
| Primary safety outcome n(%) | 13 (19.7) | 15 (21.7) | 0.98 (0.88, 1.09) | 0.70 |
| Intracranial hemorrhage n(%) | 4 (6.1) | 10 (14.5) | 0.92 (0.84, 1.01) | 0.06 |
| Malignant brain edema n(%) | 5 (7.6) | 7 (10.1) | 0.97 (0.89, 1.06) | 0.52 |
| Death n(%) | 8 (12.1) | 9 (13) | 0.99 (0.89, 1.09) | 0.84 |
| mRS 0-1 at 90 days n(%) | 27 (40.9) | 21 (30.4) | 1.2 (1.03, 1.39) | 0.02 |
| mRS 0-2 at 90 days n(%) | 39 (59.1) | 39 (56.5) | 1.05 (0.96, 1.16) | 0.30 |
| mRS 0-3 at 90 days n(%) | 44 (66.7) | 67 (68.1) | 1.01 (0.92, 1.11) | 0.792 |
| Change between baseline and D3 NIHSS median(IQR) | -8 (-12, -2) | -4 (-9, -1) | | 0.09 |
| Final infarct size (mL) median(IQR) | 16.9 (5.4, 81.1) | 21.8 (10.8, 39.9) | | 0.45 |
| Blood glucose on day 3 (mmol/L) mean(sd) | 7 (2.6) | 7.8 (3) | | 0.13 |
| Change between baseline and D3 glucose (mmol/L) mean(sd) | 0.1 (1.6) | 0.1 (3.1) | | 0.83 |

**Abbreviation:** RR: risk ratio**,** mRS: modified Rankin scale, NIHSS: National Institute of Health Stroke Scale

*N.B. Intracranial hemorrhage is defined as Heidelberg bleeding classification class 2 or above.*

**Table S7.** **Complete case analysis (no-IVT stratum)**

| Parameter | Semaglutide (n=33) | Standard Therapy (n=34) | RR (95% CI) | p-value |
|---|---|---|---|---|
| Primary safety outcome n(%) | 6 (18.2) | 10 (29.4) | 0.89 (0.76, 1.04) | 0.14 |
| Intracranial hemorrhage n(%) | 3 (9.1) | 6 (17.6) | 0.91 (0.79, 1.05) | 0.18 |
| Malignant brain edema n(%) | 2 (6.1) | 5 (14.7) | 0.91 (0.8, 1.03) | 0.14 |
| Death n(%) | 2 (6.1) | 8 (23.5) | 0.84 (0.73, 0.97) | 0.02 |
| mRS 0-1 at 90 days n(%) | 15 (45.5) | 9 (26.5) | 1.21 (1.05, 1.39) | 0.01 |
| mRS 0-2 at 90 days n(%) | 22 (66.7) | 15 (44.1) | 1.19 (1.04, 1.37) | 0.01 |
| mRS 0-3 at 90 days n(%) | 25 (75.8) | 19 (55.9) | 1.16 (1.01, 1.32) | 0.03 |
| Change between baseline and D3 NIHSS median(IQR) | -8 (-12, -4) | -3.5 (-6, 0.8) | | <0.001 |
| Final infarct size (mL) median(IQR) | 13.1 (6.2, 38.7) | 17.1 (9.8, 25.9) | | 0.44 |
| Blood glucose on day 3 (mmol/L) mean(sd) | 6.4 (1.4) | 7.8 (3.1) | | 0.02 |
| Change between baseline and D3 glucose (mmol/L) mean(sd) | -0.5 (1.1) | 0.2 (2.7) | | 0.23 |

**Abbreviation:** RR: risk ratio**,** mRS: modified Rankin scale, NIHSS: National Institute of Health Stroke Scale

*N.B. Intracranial hemorrhage is defined as Heidelberg bleeding classification class 2 or above.*

**Table S8.** **Complete case analysis (IVT stratum)**

| Parameters | Semaglutide (n=33) | Standard Therapy (n=35) | RR (95% CI) | p-value |
|---|---|---|---|---|
| Primary safety outcome n(%) | 7 (21.2) | 5 (14.3) | 1.07 (0.94, 1.23) | 0.30 |
| Intracranial hemorrhage n(%) | 1 (3) | 4 (11.4) | 0.93 (0.83, 1.03) | 0.16 |
| Malignant brain edema n(%) | 3 (9.1) | 2 (5.7) | 1.03 (0.92, 1.16) | 0.59 |
| Death n(%) | 6 (18.2) | 1 (2.9) | 1.15 (0.98, 1.3) | 0.13 |
| mRS 0-1 at 90 days n(%) | 12 (36.4) | 12 (34.3) | 1.05 (0.9, 1.22) | 0.54 |
| mRS 0-2 at 90 days n(%) | 17 (51.5) | 24 (68.6) | 0.95 (0.84, 1.06) | 0.36 |
| mRS 0-3 at 90 days n(%) | 19 (57.6) | 28 (80) | 0.89 (0.79, 1.01) | 0.06 |
| Change between baseline and D3 NIHSS median(IQR) | -8 (-12, 0) | -5 (-11, -2.5) | | 0.72 |
| Final infarct size (mL) median(IQR) | 27.7 (5.3, 127.6) | 28.6 (12.4, 59.1) | | 0.19 |
| Blood glucose on day 3 (mmol/L) mean(sd) | 7.6 (3.3) | 7.8 (2.9) | | 0.86 |
| Change between baseline and D3 glucose (mmol/L) mean(sd) | 0.7 (1.8) | -0.1 (3.4) | | 0.23 |

**Abbreviation:** RR: risk ratio**,** mRS: modified Rankin scale, NIHSS: National Institute of Health Stroke Scale

*N.B. Intracranial hemorrhage is defined as Heidelberg bleeding classification class 2 or above.*

**Supplementary Note 1: Summary of Protocol and Statistical Analysis Plan Changes**

**The following is a list of main protocol changes from protocol version 1.0 dated 16 June 2023 to version 2.0 dated 15 May 2024. The main reasons for the protocol changes are as follows:**

| Section | Protocol version 1.0 change from | Protocol version 2.0 change to | Rationale |
|---|---|---|---|
| Methods (Outcomes) | The primary outcome is good functional outcome, defined as modified Rankin Scale 0-3 at 90 days. | The primary outcome is good functional outcome, defined as modified Rankin Scale 0-2 at 90 days. Secondary outcomes include modified Rankin Scale 0-1, 0-3, and ordinal shift at 90 days. | Most clinical trials adopted mRS 0 to 2 as the definition for good neurological recovery for anterior circulation large vessel occlusion (1-3). |
| Methods (Statistical Analysis) | Multivariable logistic regression for binary outcomes. d | Multivariable modified Poisson regression for binary outcomes. | Compared to logistic regression, modified Poisson regression could provide unbiased estimation of risk ratios in the setting of common event occurrence (4). |
