## Supplementary material for "Glucagon-Like Peptide-1 Receptor Agonist in Large Vessel Occlusion Treated by Reperfusion Therapy: A Phase 2 Randomized Trial": Study protocol

### Clinical Study Protocol

#### (GALLOP study)

### 1. BACKGROUND

Endovascular thrombectomy (EVT) is a highly effective therapy for acute ischemic stroke with large vessel occlusion (LVO) [1]. Over the past decade, EVT was proven efficacious in selected patients with symptoms onset or last-known-well time of up to 24 hours [2]. With a number-needed-to-treat (NNT) of 2.3-2.8 to achieve functional independence [1, 2], EVT had become the current state-of-the-art treatment for ischemic stroke with LVO [3]. Nevertheless, more than half of LVO strokes suffered from functional dependence or death despite EVT [4].

Futile EVTs were contributed by peri-procedural malignant brain edema (MBE) and symptomatic intracranial hemorrhage (sICH) [5]. Studies suggested that 26.9% of EVTs were complicated by MBE [6], whereas sICH was present in 6-9% of LVO patients who received EVT [2, 7]. The fundamental pathophysiology of MBE and sICH is blood-brain-barrier (BBB) disruption secondary to ischemia, mechanical and reperfusion injury. These pathological processes can result in increased tissue permeability, excess production of oxygen free radicals and inflammatory response that eventually lead to hemorrhage and edema [8]. Poor collateral circulation, proximal LVOs, intravenous thrombolysis, blood pressure and glucose fluctuation had all been implicated to in MBE and sICH [6, 8]. However, these risk factors were either unmodifiable or not shown to improve EVT outcomes. The preliminary results of a recent randomized trial even suggested harmful effects of intensive blood pressure following EVT [9]. With indications of EVT are expanding to patients with prolonged ischemia and large ischemic cores [2, 10], enhancing BBB and neuronal tolerance to ischemia and reperfusion therapies may hugely impact on EVT outcomes. Recent animal models have shown that glucagon-like peptide peptide-1 receptor agonists (GLP-1RA) significantly reduced infarct volume and neurological deficits following temporary or permanent middle cerebral artery occlusion. These effects were likely due to the anti-oxidant, anti-inflammatory and anti-apoptotic properties of GLP-1RA that protected BBB integrity and ischemic neurons during induced LVO and/or reperfusion.

To date, no human studies have substantiated these postulations. We hypothesize that compared to standard reperfusion strategies, administration of GLP-1RA in LVO patients who receive EVT may prevent the development of MBE and sICH, thus improving neurological outcomes. In this randomized, open-label pilot study, we aim to determine the effect of semaglutide, a GLP-1RA, on the radiological and clinical outcomes in LVO patients undergoing EVT.

### 2. METHODOLOGY

#### 2.1. *Study overview*

In this multicenter, randomized, open-label pilot study, we aim to recruit 140 patients with LVO strokes in the terminal internal carotid artery (ICA) or proximal middle cerebral artery (MCA) who were eligible for EVT with a last-known-well (LKW) to puncture ≤ 12 hours. Patients will be randomized in a 1:1 ratio to semaglutide or standard therapy. Patients in the semaglutide group will receive the medication on the day of (D0) and 1 week (D7) after EVT. Interval imaging and blood tests will be arranged to ascertain the degree of

BBB leakage, final infarct size, inflammation and gene expression pre and post treatment (Figure 1). We shall recruit 40 patients from the Prince of Wales Hospital and 100 patients from Linyi People's Hospital.

2.2. *Inclusion criteria*

1. LVO stroke at terminal ICA or proximal M1 eligible for emergency endovascular treatment as per current treatment guideline [3].
2. LKW-to-puncture time ≤ 12 hours.
3. Age 18 years or greater.
4. National Institute of Health Stroke Scale (NIHSS) ≥ 10
5. LVO stroke due to thromboembolism or intracranial stenosis (acute or acute on chronic occlusion).
6. Patients who received computer tomographic angiography and perfusion (CTA+P).
7. Pre-stroke (24 hours prior to stroke onset) independent functional status with modified Rankin Scale (mRS) ≤ 2
8. Consent process completed as per national laws and regulation and the applicable ethics committee requirements.

2.3. *Exclusion Criteria:*

1. ASPECT score ≤ 5
2. Intracranial hemorrhage on pre-EVT imaging
3. EVT completed before randomization
4. LVO etiologies other than thromboembolism or intracranial stenosis (acute or acute on chronic total occlusion), e.g. arterial dissection, infective endocarditis on initial diagnostic imaging.
5. Estimated or known body mass index < 18 kg/$m^2$
6. Pregnancy/Lactation; female, with positive urine or serum beta human chorionic gonadotropin (β-hCG) test, or breastfeeding.
7. Creatinine clearance < 30mL/min
8. Severe or fatal comorbid illness, e.g. terminal malignancy
9. Participation in another clinical trial investigating a drug, medical device, or a medical procedure in the 30 days preceding trial inclusion.
10. History of allergy to GLP-1RA
11. Family or personal history of multiple endocrine neoplasia, medullary thyroid carcinoma, pancreatic carcinoma, known proliferative diabetic retinopathy
12. Active sepsis on randomization
13. Patients with hypoglycaemia on presentation. Defined as capillary or serum glucose level of <4mmol/L.
14. Patients prone to severe hypoglycaemia, including chronic kidney disease of estimated glomerular filtration rate of 50ml/min/1.73m^2; also those

with chronic liver disease with Child's Pugh score C or above; patients with recurrent unexplained hypoglycemia.

15. Patients already on GLP-1RA prior to screening.

16. Contraindications to iodine-based CT contrast.

*2.4. Study Procedure (Figure 1)*

1. LVO stroke patients will have received CTA and perfusion prior to screening.
2. Informed consent from patients or next of kin will be obtained for eligible patients.
3. After informed consent, patients will be randomized into semaglutide or standard treatment in a 1:1 ratio by computer-generated codes.
4. Patients randomized into the semaglutide group will receive 0.5mg subcutaneous injection of the drug before or during EVT, and 7 days after the procedure. i.e. semaglutide group will receive a total of 2 injections.
5. All study subjects will receive plain CT brain and perfusion D4-7 post EVT to look for MBE, sICH and hyperperfusion. Additional brain imaging may also be arranged as per clinical needs.
6. All study subjects will receive a standardized stroke protocol MRI D14-21 after EVT for quantification of infarct volume.
7. NIHSS before and immediately after, D3, D14-21, D90±7 post-EVT will be assessed.
8. mRS before, D14-21, D90±7 post-EVT will be assessed.
9. Blood test before and immediately, 3 days and 14 days after EVT (D0pre, D0post, D3, D14-21) will be collected for neurovascular inflammatory markers and transcriptomic analysis.
10. Capillary blood glucose, blood pressure and pulse will be measured four times daily in accordance to the standardized post-EVT protocol during the first 5 days hospitalization. The frequency of monitoring may increase according to the clinical needs.
11. The following data will be collected:
    a. Demographic data: date of birth, date of death (if applicable), smoking and drinking status
    b. Medical comorbidities: Hypertension, diabetes mellitus, hyperlipidemia, congestive heart failure, atrial fibrillation, ischemic heart disease, history of ischemic or hemorrhagic stroke, etc.
    c. Co-medications: Anticoagulants (apixaban, dabigatran, edoxaban, rivaroxaban, heparin, warfarin), antiplatelet agents (aspirin, clopidogrel, ticagrelor, cilostazol), lipid-lowering agents (simvastatin, atorvastatin, rosuvastatin, pravastatin, fluvastatin, ezetimibe, gemfibrozil, fenofibrate, erenumab), antihypertensive (angiotensin converting enzyme inhibitors, angiotensin receptor blockers, beta blockers, calcium channel blockers, diuretics, aldosterone antagonists, nitrates, etc.), non-steroidal anti-inflammatory agents or cyclo-oxygenase2 inhibitors (indomethacin, ibuprofen, diclofenac, celecoxib, etorixocib), glucose lowering drugs (metformin, gliclazide, glimepiride, empagliflozin, dapagliflozin, insulin).
    d. Routine blood tests including hemoglobin, white cell count, lymphocyte count, neutrophil count, creatinine, alanine transferase,

alkaline phosphatase, bilirubin, low-density lipoprotein cholesterol, high-density lipoprotein cholesterol, total cholesterol, triglyceride, fasting glucose, glycated hemoglobin A1c, etc. These blood tests are part of routine clinical care pre- and post-stroke.

e. Imaging data: ASPECT score [11], site of occlusion, collateral score, volume of infarct core, penumbra, mismatch volume and mismatch ratio.
f. Stroke time metrics: LKW-to-hospital, -imaging, -needle, -puncture, -reperfusion time.
g. EVT outcomes: modified thrombolysis in cerebral infarction (TICI) score [12].
h. Occurrence of MBE, asymptomatic or symptomatic ICH, hemorrhagic transformation, subarachnoid hemorrhage (see Endpoint measurement for details).

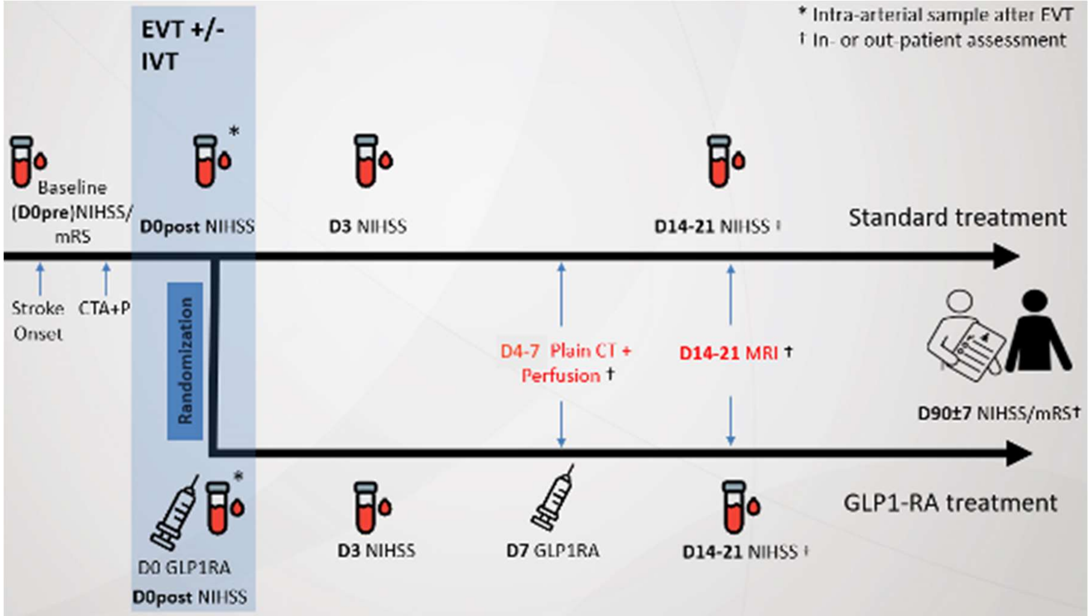


**Figure 1. Study flow diagram.**

2.5. *Endpoint measurement*

Primary efficacy outcome is D90 mRS, mRS 0-2 is considered good outcome, while mRS 3-6 is considered poor outcome.

Primary safety outcome is a composite of death, MBE and ICH.

Secondary outcomes are mRS 0-1, mRS 0-3, ordinal shift in mRS, infarct size, MBE, ICH, BBB permeability, D0post, D3, D14-21, D90±7 NIHSS, D14 mRS and death:

1. MBE is defined as [6]:
   a. Parenchymal hypodensity of at least 50% of the MCA territory and signs of local brain swelling such as sulcal effacement and compression of the lateral ventricle, AND
   b. Midline shift of ≥5 mm at the septum pellucidum or pineal gland with obliteration of the basal cisterns.
2. Symptomatic ICH is defined as any parenchymal hemorrhage or hemorrhagic transformation temporally related to any worsening in neurological condition [13].

3. ICH is defined as Heidelberg Bleeding Classification class 2 or above [14].
4. BBB permeability by CT perfusion scan
5. Certified personnel will obtain clinical outcomes as stated above. All personnel involved in determination of clinical outcomes will be blinded from treatment allocation and radiological outcomes.
6. Experienced neuroradiologists and vascular neurologists shall analysis the neuroimages as stated above. All personnel involved in imaging interpretation will be blinded from treatment allocation and the clinical outcomes.

#### 2.6. Data analysis and management

##### 2.6.1. *Magnetic Resonance Imaging*

We shall perform cranial MRI examinations in a 3-Tesla scanner (MAGNETOM Prisma; Siemens AG, Munich, Germany). The scanning protocol will employ a standard stroke protocol MRI scan with 3D T1-weighted, T2-weighted, FLAIR, susceptibility weighted imaging, diffusion weighted imaging, time-of-flight magnetic resonance angiography (MRA).

##### 2.6.2. *CT perfusion*

CT scans will be performed using 64 multidetector row scans. CT perfusion scans will be acquired in axial mode, covering the whole brain and obtained at 70kV and 200mA at 0.28sec rotation. Intravenous contrast (45mL Omnipaque 350) is administered at a rate of 5mL/second, and after a lag time of 7 seconds, serial images of the brain are acquired every 1.5 second for 15 cycles and then every 3.0 seconds for 10 cycles. The total scanning time is 52 seconds.

##### 2.6.3. *Blood test*

Approximately 10-15mL of blood will be taken at D0pre, D0post, D3 and D14-21. Samples will be stored for future analysis of gene expression and neurovascular inflammatory markers.

#### 2.7. Sample size estimation and statistical analysis

The current study will adopt a two-arm parallel group trial design in which participants will be randomly assigned to one of the two arms in a 1:1 sequence. When treating D90 mRS 0-2 as a binary outcome variable, a sample size of 120 for the phase 2 trial is recommended in order to guard against the lack of precision by using inflated estimates [15]. When treating D90 mRS on a continuous scale, the non-central t-distribution (NCT) approach rather than traditional standard sample size calculation was chosen to calculate the sample size based on two main reasons [16]. First, the variance in the traditional standard sample size calculation formula corresponds to the population variance, whereas in practice the estimated sample variance is used in calculating the sample size. This deficiency of not accounting for uncertainty in variance estimation may lead to underestimation of sample size and thus result in reduction of power, whereas the NCT approach uses cumulative distribution function to adjust for the imprecision of variance estimation [17]. Second, compared to other sample calculations with adjustment, such as the 80% and 95% upper confidence limit methods, adopting the NCT method was shown to have a lower sample size while ensuring the average anticipated power under the same design conditions of the current study. If considered NCT approach, fixed the type I and type II errors at 5% and 10%, respectively, and set the expected standardized differences in between 0.2 to 0.25, the optimal number of participants per arm for the pilot study will between 23 (for reaching 0.25 standardized difference) to 28 (for reaching 0.2 standardized difference). The optimal sample size per arm for the main trial will be 358 to 552 (90% power and 0.2 to 0.25 standardized difference). [17]. Considering approximately 2.5% suboptimal scan qualities, 5% of loss-to-follow-up and 10% of suboptimal recanalization, 140 participants are required for the phase 2 study. Sample size for the main trial will be recalculated once the estimated standardized difference is obtained from the pilot study.

Permuted blocked randomization will be employed in the current study, as it as it has the advantage of ensuring overall balance of subjects throughout trial setting [18]. An interim analysis will be performed to evaluate the safety of semaglutide treatment when at least 70 patients have completed the study. Considering that an interim analysis will be performed, smaller block sizes (i.e., size = 2, 4, and 6) will be adopted in order to avoid possible mid-block inequality caused by using larger blocks. To mask the randomization pattern, the block sizes will be randomly generated and distributed in the randomization list and a block with unbalanced treatment distribution will be generated at the start and middle of the list. The randomization process will be performed using *blockrand* package in R Studio (RStudio Team, 2022).

In the descriptive analysis, we shall express continuous variables as means (+/- standard deviation or interquartile range), and categorical data as number (percentage). We will compare baseline continuous variables of semaglutide versus standard therapy by independent sample t-test or Wilcoxon rank-sum test, and categorical variables by Chi-squared or Fisher's exact test as appropriate.

Primary analysis will be performed based on the intention-to-treat principle, i.e. all patients who underwent randomization. Multivariable Poisson regression will be used to compare the binary (primary and secondary) study endpoints except death, with the primary effect parameter being the risk ratio with 95% confidence interval. Cox proportional-hazards model will be used to compare death, hazard ratio with 95% confidence interval will be reported. Ordinal logistic regression will be used to compare the ordinal shift in mRS across the two treatment arms after checking the proportional-odds assumption by Brant test. Continuous endpoints will be compared by analysis of covariance (ANCOVA). Comparisons of the primary outcome (mRS 0-2 at 90 days), ordinal shift of mRS, mRS 0-3 and mRS 0-1 at 90 days will be adjusted for a pre-specified set of covariates, including age, premorbid mRS, intravenous thrombolysis status, NIHSS on presentation, LKW-to-puncture time, mTICI, and baseline ASPECTS. Comparisons of other secondary outcomes will be adjusted for NIHSS on presentation only due to the anticipated low number of events. Missing outcomes were imputed as worse possible score for an outcome measure (i.e. mRS 6; NIHSS 42).

Per-protocol sample includes participants with no clinically meaningful deviation from the protocol, such as not being in line with inclusion criteria; and who have completed the study successfully. Participants not being able to receive the second dose of semaglutide due to critical medical conditions or death will not be excluded from the per-protocol sample. Per-protocol analyses will be conducted to compare the primary and secondary endpoints.

##### 2.8. *Research subject protection and ethical issues*

Patients in the semaglutide arm will receive 2 doses of the 0.5mg subcutaneous injection of the medication (see below). Furthermore, all patients will receive additional blood tests and CT perfusion scan in addition to routine care. We shall protect the patient by:

a) Minimizing venipuncture by combining blood tests with those required for routine clinical care.
b) Lowest possible radiation dose is adopted for all CT protocols.
c) We shall not proceed with imaging if patient is clinically unstable for transfer.
d) Capillary blood glucose monitoring in the first 5 days of study or longer if deemed clinically necessary. Blood glucose monitoring is part of the routine post-EVT care in PWH and Linyi People's Hospital. Most of the stroke patients who received EVT will be kept nil per os for 12-24 hours with capillary blood glucose monitoring every 4 hours. During the period, diabetic medications will not be administered unless the glucose level is > 10mmol/L. Insulin and oral hypoglycemic agent will then be resumed gradually after reviewing the blood glucose level to maintain euglycemia after the resumption of diet.
e) Exclude patients with known iodine-based contrast allergy.
f) Exclusion of patients with history of hypersensitivity reaction to GLP-1RA, history of or active malignancy, family history of multiple endocrine neoplasia, proliferative retinopathy, pancreatic pathologies or underweight with BMI <18.5kg/m$^2$;
g) Exclusion of patients with history of hypersensitivity reaction to gadolinium-based contrast and chronic kidney disease KDIGO stage 4 or above.

#### 3. **INVESTIGATIONAL PRODUCT**

The drug under the investigation of this study is called semaglutide. The dosage frequency is 0.5mg once weekly via subcutaneous injection twice for this study. As GLP-1R agonist mainly facilitates post prandial insulin release, hypoglycemic complications are rare. We shall not adjust the usual diabetic treatment unless patients have pre-existing hypoglycemic episodes suggested by capillary blood glucose monitoring or neuroglycopenic symptoms. For non-diabetic patients, GLP-1R agonist use, according to literature review, does not cause hypoglycemia in the absence of other hypoglycemic agents.

Side effects of GLP-1RA, including nausea (17.0%-19.9%), diarrhea (12.2%-13.3%), vomiting (6.4-8.4%), pancreatitis, hemorrhagic retinopathy, drug hypersensitivity, malignant neoplasms of thyroid (frequency not defined), have been reported [19]. We anticipate a very low adverse event rate as only 2 injections will be given to the GLP-1RA arm.

A semaglutide dosage of 0.5mg was chosen based on the 12µg/kg dosing in an rat model that evaluated the effect of semaglutide with induced MCA occlusion followed by reperfusion [20]. A 2µg/kg dosing was estimated after conversion of animal dose to human equivalent dose by a factor of 0.162 under the FDA guidelines. Since the available dosages of semaglutide are 0.25mg, 0.5mg, 1mg and 2mg, 0.5mg per injection is a reasonable heuristic dosage.

***Safety Assessment***

The following measures will be performed to ensure subjects are suitable to proceed study procedures during the study period:

a. Exclusion of patients with contraindication to GLP-1RA (details see ***Exclusion Criteria***)
b. We shall perform capillary blood glucose monitoring for participants during the hospitalization 4 times daily during the first 5 days of study if the patient has not been discharged. The duration and frequency of monitoring may increase according to clinical needs.
c. We shall also exclude patients with hypoglycaemia on presentation, defined as capillary or serum glucose level of <4mmol/L.

#### 4. ADVERSE EVENTS

The investigators are responsible for the detection and documentation of events meeting the criteria and definition of an adverse events (AE) or severe adverse events (SAE), as provided in this protocol. During the study when there is a safety evaluation, the investigators or site staff will be responsible for detecting, documenting and reporting AEs and SAEs. SAEs will be reported to ethics committee.

##### 4.1. *Definition of AE*

Any untoward medical occurrence in a patient or clinical investigation subject, temporary associated with the use of a medicinal product, whether or not considered related to the medicinal product. An AE can therefore be any unfavorable and unintended sign (including an abnormal laboratory finding), symptom, or disease (new or exacerbated) temporally associated with the use of a medicinal product. For marketed medicinal products, this also includes failure to produce expected benefits (*i.e.* lack of efficacy), abuse or misuse

*Examples of an AE* ***include****:*

- Significant or unexpected worsening or exacerbation of the condition/indication under study. ("Lack of efficacy" per se will not be reported as an AE. The signs and symptoms or clinical sequelae resulting from lack of efficacy will be reported if they fulfill the AE or SAE definition (including clarifications).)
- Exacerbation of a chronic or intermittent pre-existing condition including

either an increase in frequency and/or intensity of the condition.

- New conditions detected or diagnosed after investigational product administration even though it may have been present prior to the start of the study.

- Signs, symptoms, or the clinical sequelae of a suspected interaction

*Examples of an AE* ***do not include*** *a/an:*

- Medical or surgical procedure (*e.g*., endoscopy, appendectomy); the condition that leads to the procedure is an AE.

- Situations where an untoward medical occurrence did not occur (social and/or convenience admission to a hospital).

- Anticipated day-to-day fluctuations of pre-existing disease(s) or condition(s) present or detected at the start of the study that do not worsen.

- The disease/disorder being studied, or expected progression, signs, or symptoms of the disease/disorder being studied, unless more severe than expected for the subject's condition

##### *4.2.* ***Definition of SAE***

An SAE is any untoward medical occurrence that, at any dose:

- *Results in death*

- *Is life-threatening* (NOTE: The term 'life-threatening' in the definition of 'serious' refers to an event in which the subject was at risk of death at the time of the event. It does not refer to an event, which hypothetically might have caused death, if it were more severe.

- *Requires hospitalization or prolongation of existing hospitalization*; NOTE: In general, hospitalization signifies that the subject has been detained (usually involving at least an overnight stay) at the hospital or emergency ward for observation and/or treatment that would not have been appropriate in the physician's office or out-patient setting. Complications that occur during hospitalization are AEs. If a complication prolongs hospitalization or fulfils any other serious criteria, the event is serious. When in doubt as to whether "hospitalization" occurred or was necessary, the AE should be considered serious. Hospitalization for elective treatment of a pre-existing condition that did not worsen from baseline is not considered an AE.

- Results in disability/incapacity
- Is a congenital anomaly/birth defect

#### 5. ACCOUNTABILITY AND COMPLIANCE

##### *5.1. **Product Accountability***

The investigator and staff will maintain investigational product accountability records throughout the course of the study. The responsible person(s) will document the amount of investigational product dispensed and the amount supplied and/or administered to and returned by subjects, if applicable.

##### 5.2. ***Withdrawal Criteria***

Subjects may withdraw from study at any time and for any reason. The investigator (or designee) must document the reason for withdrawal. The investigator may also withdraw subjects for the following reasons:

- Adverse events (AE) or Serious Adverse Events (SAE), both related or not related to study drug that cause subject no longer suitable to participate study
- Lost to follow up
- Subject decide to withdraw from the study

#### 6. GOOD CLINICAL PRACTICE AND DECLARATION OF HELSINKI

This study will be performed in compliance with the International Council for Harmonisation of Technical Requirements for Pharmaceuticals for Human Use (ICH) Good Clinical Practice (GCP) and the Declaration of Helsinki. It is designed based on a thorough knowledge of the scientific background, a careful assessment of risks and benefits, a reasonable likelihood of benefit to the population studied and will be conducted by suitably trained investigators using approved protocols.

#### GALLOP Study - Schedule of Study Activities

| | **Screening** (LKW-to-puncture time ≤ 12 hours) | **Randomization** | **Treatment** (Before or during EVT) | | | | | | **Imaging** | **Treatment** | **Imaging** | **Follow-up** |
|---|---|---|---|---|---|---|---|---|---|---|---|---|
| **Study Visit** | | **D0pre** | | **D0post** | | | **D3** | | **D4-7** | **D7** | **D14-21** | **D90±7** |
| **Day** | **0** | **0** | **0** | **0** | **1** | **2** | **3** | **4** | **4-7** | **7** | **14-21** | **90±7** |
| Informed consent | X | | | | | | | | | | | |
| Demographic | X | | | | | | | | | | | |
| Body weight | | | | | | | | | | | | |
| Medical comorbidities | X | | | | | | | | | | | |
| Co-medications | X | | | | | | | | | | | |
| Inclusion/exclusion criteria | X | | | | | | | | | | | |
| Enrollment (randomization) | | X | | | | | | | | | | |
| EVT | | | X | | | | | | | | | |
| CTA+P | X | | | | | | | | | | | |
| CTB + perfusion | | | | | | | | | X | | | |
| MRI (stroke protocol) | | | | | | | | | | | X | |
| NIHSS | | X | | X | | | X | | | | X | X |
| mRS | | X | | | | | | | | | X | X |
| CBG[a] | | X | | X | X | X | X | X | | | | |
| BP/P[a] | | X | | X | X | X | X | X | | | | |
| Routine blood test[b] | X | | | X | | | X | | | | X | X |
| Biomarker sample collection[c] | | X | | X | | | X | | | | X | |
| [IMP] GLP-1RA | | | X | | | | | | | X | | |

a. Capillary blood glucose, blood pressure and pulse will be measured at least 4 times daily during the first 5 days of study.
b. Routine blood tests including hemoglobin, white cell count, lymphocyte count, neutrophil count, creatinine, alanine transferase, alkaline phosphatase, bilirubin, low-density lipoprotein cholesterol, high-density lipoprotein cholesterol, total cholesterol, triglyceride, fasting glucose, glycated hemoglobin A1c, etc.
c. Biomarker sample collection for neuroinflammatory markers and transcriptomic analysis. Blood will be collected before and immediately, 3 days and 14 days after EVT (D0pre, D0post, D3, D14-21).
